# Analysis of rare coding variants in 470,000 UK Biobank participants reveals genetic associations with childhood asthma predisposition

**DOI:** 10.1101/2025.02.12.25322158

**Authors:** Zhenzhen Liu, David Curtis

## Abstract

**Background:** Previous studies of genetic contributions to risk of childhood asthma have implicated common variants with small effect sizes. Some studies using exome sequence data have reported associations with rare coding variants having larger effects on risk, notably an analysis of 145,000 subjects which found association with loss of function (LOF) variants in *FLG*, the gene coding for filaggrin. Here we report the results of an analysis of rare nonsynonymous and LOF variants, carried out on the full UK Biobank cohort of 470,000 exome-sequenced participants.

**Methods:** The phenotype of childhood asthma was defined as reporting asthma with onset before 18. Regression analysis was applied to gene-wise tests for association LOF and nonsynonymous variants. 45 tests using different pathogenicity predictors were applied to the first cohort of 200,000 participants. Subsequently the 100 genes showing strongest evidence for association were analysed in the second cohort of 270,000 participants, using only the best-performing predictor for each gene. For *FLG*, separate analyses were carried out for participants with atopic dermatitis.

**Results:** Three genes achieved statistical significance, *FLG, IL33* and *PRKCQ*. The effects on asthma risk and frequencies of variants in different functional categories were characterised for these genes. Damaging coding variants were associated with increased risk of asthma in *FLG* and *IL33* but reduced risk in *PRKCQ. FLG* LOF variants were also associated with risk of atopic dermatitis and their effect on asthma risk was higher in people who reported a diagnosis of atopic dermatitis.

**Conclusions:** Rare coding variants in a small number of genes have important effects on asthma risk. Further study of individual variant effects might elucidate mechanisms of pathogenesis.

This research has been conducted using the UK Biobank Resource.

## Introduction

Asthma is a chronic inflammatory disease in which environmental risk factors are important but which has high heritability, with estimates ranging from 35% to 95% (Ober and Yao, 2011; Pijnenburg *et al*., 2022). Asthma can be broadly divided into two endotypes: the T2-high endotype, which is associated with early-onset, allergic asthma, and the T2-low endotype, which is associated with late-onset, non-allergic asthma (Sansone *et al*., 2020). The heritability of early-onset asthma is particularly high, at 68% to 92% (Ma *et al*., 2020).

Genome-wide association studies (GWASs) have implicated hundreds of common variant associations with asthma traits (Espuela-Ortiz et al., 2023). Genes in implicated regions are involved in inflammatory and secondary immune responses, including B-cell and T-cell receptors, immunoglobulin E levels and eosinophil counts, while other studies report genes associated with obesity, mucus secretion, the PI3K/AKT pathway and calcium ion transport (Han *et al*., 2020; Zhu *et al*., 2021; Espuela-Ortiz *et al*., 2023).

Variants identified through GWAS are associated with small effect sizes and it can be difficult to determine their functional effects whereas rarer variants directly impacting coding regions of genes can have larger and more readily interpretable effects. Two low frequency loss of function (LOF) variants, denoted R501X and 2282del4, in *FLG*, the gene for filaggrin, increase the risk of ichthyosis vulgaris and atopic dermatitis in populations of European ancestry and increase the risk of asthma in children who have atopic dermatitis (Marenholz *et al*., 2006; Smith *et al*., 2006). A subsequent meta-analysis found that LOF *FLG* variants were associated with asthma only in children with a prior diagnosis of atopic dermatitis (Rodriguez *et al*., 2009). A preprint presenting a study of exome sequence data from 24,576 asthma cases and 120,530 controls from the Scandinavian Asthma Genetic Study and UK Biobank also reports that rare LOF in *FLG* are collectively associated with increased childhood asthma risk (Cameron-Christie A. Mackay Q. Wang H. Olsson B. Angermann G. Lassi J. Lindgren M. Hühn Y. Ohne M. Gavala J. Wang G. Povysil S. V. V. Deevi G. Belfield I. Dillmann D. Muthas S. Cohen S. Young A. Platt S. Petrovski, 2020). An early sequencing study of candidate genes claimed that rare, non-coding variants in *IL12RB1* were associated with asthma but this finding has not been replicated (Torgerson *et al*., 2012). A study of rare and low frequency coding variants using array genotypes implicated gene-wise association with functional variants in *GSDMB* significant after correction for multiple testing (Igartua *et al*., 2015). However the authors report that this signal was mostly driven by rs12450091in the Mexican American subset of their data and since this variant has a high frequency in Latinos (minor allele frequency (MAF) 0.19) there is a possibility that this finding may be related to the previously identified common variant association at this locus. Whole-genome sequencing and imputation into the Icelandic population led to the identification of a rare canonical splice site variant, rs146597587-C, in the GWAS-implicated gene *IL33* which is associated with lower eosinophil counts and reduced risk of asthma in Europeans (Smith *et al*., 2017).

Exome sequence data has now been made available for 470,000 UK Biobank participants, allowing a more thorough exploration of the effects of rare coding variants on childhood asthma in the general population.

## Methods

The exome sequence data for UK Biobank participants was released in two phases. Data had been downloaded for 200,632 subjects who had undergone exome-sequencing and genotyping by the UK Biobank Exome Sequencing Consortium using the GRCh38 assembly with coverage 20X at 95.6% of sites on average (Szustakowski *et al*., 2021). Subsequently, the UK Biobank Research Analysis Platform (RAP) made available the Final Release Population level exome variants in PLINK format for 469,818 exomes which had been produced at the Regeneron Genetics Center based on DNA extracted from stored blood samples and using the protocols described here: https://dnanexus.gitbook.io/uk-biobank-rap/science-corner/whole-exome-sequencing-oqfe-protocol/protocol-for-processing-ukb-whole-exome-sequencing-data-sets (Backman *et al*., 2021). Data from the first cohort of 200,000 participants was used for exploratory first stage analyses and then data from the remaining 270,000 participants was used in confirmatory analyses. UK Biobank had obtained ethics approval from the North West Multi-centre Research Ethics Committee which covers the UK (approval number: 11/NW/0382) and had obtained informed consent from all participants. The UK Biobank approved an application for use of the data (ID 51119) and ethics approval for the analyses was obtained from the UCL Research Ethics Committee (11527/003).

The phenotype of childhood asthma was based on participants’ responses to the questionnaire item, “Age asthma diagnosed by doctor.” Participants who reported to be diagnosed at or before the age of 18 were categorised as cases, while the remainder served as controls. There was no effort to exclude control subjects who might have had alternative allergic disease diagnoses. Among 469,765 participants, 34,474 cases were identified. This ratio closely approximates the prevalence of childhood asthma in the British population (∼1 in 11 individuals).

Attention was restricted to variants with minor allele frequency (MAF) < 0.01. All exonic variants were annotated using Variant Effect Predictor (VEP) (McLaren *et al*., 2016). Nonsynonymous variants were additionally annotated using the AlphaMissense plug-in of VEP, which produces a raw score and a categorisation of likely pathogenic, likely benign or ambiguous, these three categories being converted to numerical scores of 2, 0 or 1 respectively (Cheng *et al*., 2023). Additionally, rank prediction scores were obtained for an additional 43 different predictors of pathogenicity as provided for all possible nonsynonymous variants in dbNSFP v4 (Liu *et al*., 2020).

To obtain population principal components reflecting ancestry, version 2.0 of *plink* (https://www.cog-genomics.org/plink/2.0/) was run with the options *--maf 0.1 --pca 20 approx* (Chang *et al*., 2015; Galinsky *et al*., 2016).

In order to carry out weighted burden analysis, for each LOF or nonsynonymous variant a score was defined. For LOF variants this score used a parabolic function of allele frequency as previously described, with extremely rare variants (MAF ∼ 0) being assigned a score of 10 and less rare variants (MAF=0.01) being assigned a score of 1 (Curtis, 2012, 2022). For nonsynonymous variants, this frequency score was assigned in the same way but was then multiplied by the score for a pathogenicity predictor in order to produce an overall score for that variant. For each gene, each participant would receive a LOF score and a nonsynonymous score, consisting of the sums of the scores for the individual variants of each class carried by that participant. To test for association, multiple logistic regression was performed using sex and 20 principal components as covariates. For the null model, asthma phenotype was predicted solely from these covariates. For the alternative model, asthma phenotype was predicted from these covariates along with the LOF and nonsynonymous scores. The likelihoods of these models were compared and a p value obtained assuming that twice the log likelihood difference followed a chi-squared distribution with two degrees of freedom and the statistical evidence for association was summarised as minus log10(p) (MLP).

Because previous work had shown that LOF variants and nonsynonymous variants could make different relative contributions to disease risk and because different predictors of pathogenicity had differential performance across different genes, a two-stage approach to analysis was followed (Curtis, 2024). In the first stage, the first cohort of 200,000 exome-sequenced participants was used to carry out 45 different regression analyses as described above for each protein-coding gene using each of the 45 pathogenicity predictors. This yielded 45 MLPs for each gene, the maximum being denoted MaxMLP. In the second stage, the remaining 270,000 participants were used for analyses of only the 100 genes showing the strongest evidence for association in the first phase and each gene was analysed using only the predictor which had produced the maximum MLP for that gene in the first phase. Thus only 100 tests were performed in the second phase, meaning that a gene could be declared statistically significant if it produced a test statistic significant at p < 0.05/100.

For each gene achieving statistical significance, the analysis was repeated using the best predictor in all 470,000 participants. In the case of *FLG*, this analysis was repeated separately in those with and without a diagnosis of atopic dermatitis, determined from having a relevant ICD9 or ICD10 code or a self-report of “eczema/dermatitis”. Additionally, a full logistic regression analysis was carried out in all participants including the same covariates and using the raw counts for all categories of variants as characterised by VEP, along with counts of variants having a dbNSFP rank score for the best performing predictor of more than a threshold value of 0.35.

## Results

In the first stage analysis, MLPs were obtained for 19,008 protein coding genes using 45 different predictors and these are listed in Supplementary Table 1. The 100 genes with the highest MaxMLPs went forward into the second stage analysis and Supplementary Table 2 shows the MLPs obtained in the second cohort of 270,000 participants using the same predictor as had yielded the maximum MLP in the first 200,000 participants. Of these, three genes reached the threshold for statistical significance, *FLG, PRKCQ* and *IL33*, and these results are shown in Table 1. As can be seen, all three of these genes produced MLPs exceeding the critical threshold of -log10(0.05/100) = 3.3 in the second stage analysis. In the full sample of 470,000, these three genes all also produced MLPs which would exceed a conservative threshold for multiple-testing of -log10(0.05/(19008*45)) = 7.23.

**Table 1.** For each gene achieving statistical significance in the second stage analysis, the table shows the maximum -log10(p) (MLP) produced by any of 45 pathogenicity predictors in the first stage analysis of 200,000 participants along with the predictor producing this MLP, results obtained using the same predictor in the second stage using 270,000 participants and in the total sample of 470,000. All results are obtained from multiple logistic regression analyses of childhood asthma with sex and 20 principal components as covariates and MAF-weighted LOF scores and pathogenicity scores as predictors.

| Gene | MaxMLP200K | Predictor | MLP270K | MLP470K |
| --- | --- | --- | --- | --- |
| *FLG* | 14.17 | AM_prediction | 15.01 | 27.65 |
| *PRKCQ* | 5.98 | AM_prediction | 3.77 | 9.28 |
| *IL33* | 4.43 | BayesDel_addAF_rankscore | 4.13 | 8.07 |

**Table 2.**
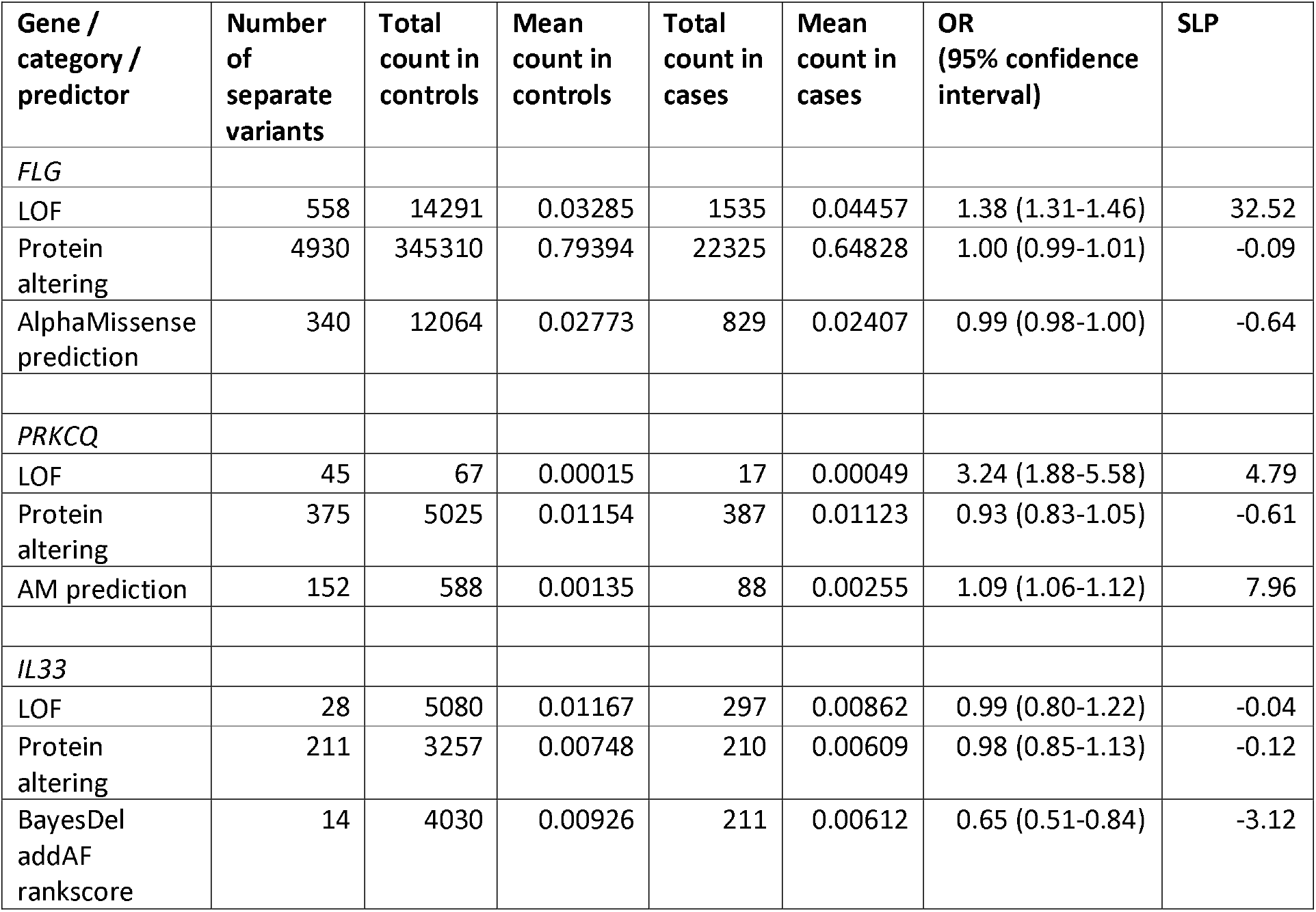
Table showing results of multiple logistic regression analysis of childhood asthma using raw counts of LOF and protein altering variants as well as those variants having a score of > 0.35 for the relevant predictor. Analyses were carried out in the full sample of 470,000 participants.

| Gene / category / predictor | Number of separate variants | Total count in controls | Mean count in controls | Total count in cases | Mean count in cases | OR (95% confidence interval) | SLP |
| --- | --- | --- | --- | --- | --- | --- | --- |
| *FLG* |  |  |  |  |  |  |  |
| LOF | 558 | 14291 | 0.03285 | 1535 | 0.04457 | 1.38 (1.31-1.46) | 32.52 |
| Protein altering | 4930 | 345310 | 0.79394 | 22325 | 0.64828 | 1.00 (0.99-1.01) | -0.09 |
| AlphaMissense prediction | 340 | 12064 | 0.02773 | 829 | 0.02407 | 0.99 (0.98-1.00) | -0.64 |
| *PRKCQ* |  |  |  |  |  |  |  |
| LOF | 45 | 67 | 0.00015 | 17 | 0.00049 | 3.24 (1.88-5.58) | 4.79 |
| Protein altering | 375 | 5025 | 0.01154 | 387 | 0.01123 | 0.93 (0.83-1.05) | -0.61 |
| AM prediction | 152 | 588 | 0.00135 | 88 | 0.00255 | 1.09 (1.06-1.12) | 7.96 |
| *IL33* |  |  |  |  |  |  |  |
| LOF | 28 | 5080 | 0.01167 | 297 | 0.00862 | 0.99 (0.80-1.22) | -0.04 |
| Protein altering | 211 | 3257 | 0.00748 | 210 | 0.00609 | 0.98 (0.85-1.13) | -0.12 |
| BayesDel addAF rankscore | 14 | 4030 | 0.00926 | 211 | 0.00612 | 0.65 (0.51-0.84) | -3.12 |

For *FLG*, results were statistically significant for both the 13,828 participants with a diagnosis of eczema and those without, with MLPs of 19.40 and 9.08, respectively. However the estimate for the effect size of LOF variants was higher in those with eczema (beta = 0.0913, SE = 0.0096) than those without (beta = 0.0249, SE = 0.0039).

Table 2 shows the results for these genes of analysing the raw counts, not weighted by frequency, of variants in LOF and protein altering categories assigned by VEP along with variants scoring higher than 0.35 with the relevant predictor. Multiple linear regression analysis to quantify the effect on childhood asthma was carried out using all variant counts along with sex and 20 principal components. Other variant categories were included in the analyses, consisting of 3’ and 5’ UTR, splice region, synonymous and intronic but no category was significant even at a nominal level of 0.05 for any gene so results for these other variant categories are not shown.

It can be seen that for *FLG* LOF variants are observed at hundreds of separate locations and are cumulatively not very rare, with an average rare variant load per participant of around 3%. (This does not include the two previously reported common LOF variants, R501X and 2282del4, which were not considered in this analysis.) On average, they exert a modest effect on risk of childhood asthma with OR 1.38. By contrast, there is no detectable effect on risk from nonsynonymous variants, even those with high scores with AlphaMissense, the best performing predictor for this gene. Detailed inspection of the results from the first cohort of 200,000 participants revealed that in fact the overall MLP obtained varied very little between predictors of nonsynonymous variant pathogenicity, consistent with the notion that the observed association was entirely due to LOF rather than nonsynonymous variants.

This situation is reversed for *IL33*, where the effect on risk is due to nonsynonymous variants with high scores for the BayesDel addAF predictor while LOF variants have no detectable effect (Feng, 2017). The nonsynonymous variants with the high scores have a protective effective, with OR 0.65. Although LOF variants are not associated with any overall effect on risk, detailed inspection of the results revealed that the essential splice site variant rs146597587-C at chr9:6255967 had MAF 0.0045 in controls and 0.0030 in controls, equivalent to 0R = 0.67, uncorrected for covariates, and consistent with the previous reports of a protective effect for this variant (Smith *et al*., 2017).

Finally, *PRKCQ* demonstrates clear signals for both LOF variants and those having high scores with AlphaMissense. While the LOF variants variants are rare, they have a larger estimated effect size than those with high AlphaMissense scores, with OR of 3.24 versus 1.09.

## Discussion

The findings from this study of a rare coding variants in a large exome-sequenced sample enlarges our understanding of the role of *FLG* and IL33 in susceptibility to childhood asthma, while the findings for PRKCQ are novel. For FLG we observe effects for hundreds of rare LOF variants, similar to those previously noted for common LOF variants and we note that nonsynonymous variants do not influence risk. For IL33 we note that although the previously reported essential splice site variant is protective, other LOF variants appear to have no effect whereas some rare nonsynonymous variants are also protective. For PRKCQ, both LOF and nonsynonymous variants increase risk.

The findings obtained for FLG are entirely consistent with what is known about its biological function. The gene expressed in the keratohyalin granules of skin keratinocytes codes for profilaggrin is a 400-kDa, highly phosphorylated, inactive polymer that contains 10-12 filaggrin repeats (Osawa, Akiyama and Shimizu, 2011). Upon being secreted into the intercellular space, it is rapidly dephosphorylated and cleaved into multiple filaggrin monomers and these contribute to the epidermal differentiation process and the barrier function of the stratum corneum of the skin. Notably, the C-terminal domain of profilaggrin, approximately 160 amino acids in length, is essential for the proper cleavage and subsequent release of filaggrin monomers (Shamilov, Robinson and Aneskievich, 2021). This means that any LOF variant within FLG will mean that it will yield no filaggrin molecules. By contrast, a single nonsynonymous variant in FLG can only affect one of the filaggrin copies it codes for. This accords with our observation that LOF variants can produce an effect on risk while nonsynonymous variants do not.

Notably, *FLG* is expressed in the outer layers of the oral and nasal mucosae but not in the epithelial cells of the bronchi and bronchioles, which are the primary tissues affected by asthma (Ying *et al*., 2006). As previously noted, LOF variants in *FLG* increase the risk of ichthyosis vulgaris and atopic dermatitis and increase the risk of asthma only in children who have atopic dermatitis (Marenholz *et al*., 2006; Smith *et al*., 2006; Rodriguez *et al*., 2009). In our own study, we note that the effect of *FLG* LOF variants on asthma risk is much larger in participants who report a diagnosis of dermatitis or eczema than those who do not. Although the effect is still highly significant in those who do not report this diagnosis, this does not in fact mean that *FLG* LOF variants can cause asthma in people who do not have atopic dermatitis because some participants might have actually had this diagnosis without reporting it. It remains plausible that *FLG* LOF variants exert their effect on asthma risk by interfering with the barrier function of the skin, allowing external allergens to enter and provoke allergic responses which can include both atopic dermatitis and asthma (Hsu, Akiyama and Shimizu, 2008).

The rare essential splice site variant in *IL33*, rs146597587-C, has been previously reported to be protective against childhood asthma and associated with lower blood eosinophil counts (Smith *et al*., 2017). *IL33* codes for interleukin-33 and common variants in it are also associated with asthma risk (Queiroz *et al*., 2017; Saikumar Jayalatha *et al*., 2021). Alternative splicing of IL33 has been reported to be associated with cytoplasmic localisation in epithelial cells and with markers of increased type 2 inflammation in asthma (Gordon *et al*., 2016). Interestingly, in our dataset we note that while rs146597587-C has a protective effect with raw OR = 0.67, overall LOF variants in IL33 have a null effect, with OR (95% CI) = 0.99 (0.80-1.22), suggesting that the effect of rs146597587-C might not be simply due to haploinsufficiency. We also note a substantial protective effect of nonsynonymous variants with high scores for the BaysDel addAF predictor, with OR (95% CI) = 0.65 (0.51-0.84). An inhibitor of interleukin-33 activity, astegolimab, which binds to its receptor has been found to reduce exacerbations in patients with severe asthma but was not effective against atopic dermatitis (Kelsen *et al*., 2021; Maurer *et al*., 2022). Our results enhance the evidence that interleukin-33 activity does have some role in asthma pathogenesis and that further attempts to perturb this activity may have therapeutic potential.

*PRKCQ* encodes the protein kinase C theta (PKCθ), a member of the protein kinase C (PKC) family, which plays a role in the activation and regulation of T cells. PKCθ is involved in T-cell receptor signalling, particularly in the formation of the immune synapse (Brezar, Tu and Seddiki, 2015). Upon recruitment to the immune synapse, PKCθ activates downstream signalling pathways to drive T-cell activation, differentiation, and immune function (Lin and Wang, 2004). (Lin and Wang, 2004). PKCθ also modulates the balance between effector T cells (Teff) and regulatory T cells (Treg), influencing the inflammatory responses in various immune-related conditions (Brezar, Tu and Seddiki, 2015). Studies in PKCθ-deficient mice have demonstrated a reduction in the numbers and activation of type 2 innate lymphoid cells (ILC2s) and TH2 cells, while pharmacological inhibition of PKCθ led to decreased airway inflammation (Madouri *et al*., 2017). Additionally, PKCθ-deficient mice showed impaired T-cell responses and reduced cytokine production (Anderson *et al*., 2006). Based on expected therapeutic applications, a range of PKCθ inhibitors have been developed (Curnock *et al*., 2014; Papa *et al*., 2021; Crosignani *et al*., 2024). However taken at face value our own findings suggest that reduced function of PKCθ might not lead to reduction in inflammatory processes, at least in the context of childhood asthma. This is because our results show an association of increased, not reduced, asthma risk for both LOF variants and those scored highly by AlphaMissense.

To conclude, this study of rare coding variants in a large exome-sequenced sample provides some further insights into the biological processes involved in susceptibility to childhood asthma. Specific functional studies of the effects of individual variants might provide further illumination and suggest novel therapeutic approaches.

## Data availability

The raw data is available on application to UK Biobank at https://ams.ukbiobank.ac.uk/ams/. Derived variables will be returned to UK Biobank, from which they will become available in due course. Software and scripts used to perform the analyses is available at https://github.com/davenomiddlenamecurtis.

## Acknowledgments

This research has been conducted using the UK Biobank Resource under Application Number 51119. The authors wish to acknowledge the staff supporting the High Performance Computing Cluster, Computer Science Department, University College London. The authors wish to thank the participants who volunteered for the UK Biobank project. This work uses data provided by patients and collected by NHS England as part of their care and support. This research also used data assets made available by National Safe Haven as part of the Data and Connectivity National Core Study, led by Health Data Research UK in partnership with the Office for National Statistics and funded by UK Research and Innovation (grants MC_PC_20029 and MC_PC_20058). For the work to carry out the analyses reported here, the authors did not receive any specific grant from funding agencies in the public, commercial, or not-for-profit sectors.

